# On the persistence of mental health deterioration during the COVID-19 pandemic by sex and ethnicity in the UK: evidence from Understanding Society

**DOI:** 10.1101/2021.08.04.21261600

**Authors:** Eugenio Proto, Climent Quintana-Domeque

## Abstract

We use longitudinal data from a representative sample of the UK and compare self-reported mental health, as measured by the GHQ-12 score, at three timepoints (2017-2019, April 2020 and March 2021), for the whole sample and by sex and ethnicity. Out of the 14,382 individuals interviewed in 2017-2019 and April 2020, 10,445 were interviewed again in March 2021. The mean GHQ-12 in April 2020 is 12.37 [95% CI: 12.22, 12.52] and in March 2021 is 12.36 [95% CI: 12.21, 12.51], above that in 2017-2019, 11.13 [95% CI: 10.99, 11.26]. We do not find evidence that the level of mental health goes back to pre-pandemic levels. In terms of inequalities, while the gender gap (mean difference between women and men) in mental health deterioration among White British is closing, there is no clear evidence that the ethnic gap (mean difference between ethnic minorities and White British) among men is changing.

## Introduction

The COVID-19 pandemic has contributed to a deterioration of the mental health across different populations, with some demographic groups more affected than others. In the UK, women^1,2,3^ and individuals from ethnic minorities^4^ have experienced higher increases in mental distress. In this short communication we investigate whether the deterioration in mental health has been persistent. We use longitudinal data from a representative sample of the UK and compare self-reported mental health at three timepoints (2017-2019, April 2020 and March 2021), for the whole sample and by sex and ethnicity.

## Methods

Participants were from Understanding Society, the UK Household Longitudinal Study (UKHLS), which is a large, national, probability-based survey. Sampling design features are accounted for (Supplement). Our main outcome of interest is the GHQ-12 score, a well-known self-report instrument for evaluating mental health, which goes from 0 (minimum) to 36 (maximum mental distress)^5^. We focus on participants interviewed in three waves: 2017-2019, April 2020 and March 2021. Out of the 14,382 individuals interviewed in 2017-2019 and April 2020, 10,445 were interviewed again in March 2021. The attrition rate between April 2020 and March 2021 was 25.7%. Men (n=6,096), ethnic minorities (n=1,299) and individuals with a higher GHQ-12 score are more likely to be attritors (Supplement).

We calculate the mean of the GHQ-12 score, for the whole sample and by sex (men and women) and ethnicity (ethnic minorities and White British), over time (2017-2019, April 2020, March 2021). Second, we run OLS regressions of the changes in mental health from 2017-2019 to April 2020, and from 2017-2019 to March 2021, on sex, ethnicity and their interaction, without and with controls variables (Supplement). Analyses were conducted with Stata (version 16.1). Inference was conducted using 2-sided P-values and p<0.05. The University of Essex Ethics Committee approved all data collection (Supplement).

## Results

The mean GHQ-12 in April 2020 is 12.37 [95% CI: 12.22, 12.52] and in March 2021 is 12.36 [95% CI: 12.21, 12.51], above that in 2017-2019, 11.13 [95% CI: 10.99, 11.26] (**Figure 1, panel A**). The means by sex and ethnicity are similar in April 2020 and March 2021 (**Figure 1, panel B**). **Figure 2** displays the regression coefficients (Supplement) capturing the average change in mental health gaps by sex (gender gap) and ethnicity without (**Figure 2, panel A**) and with controls (**Figure 2, panel B**). Both the gender gap (mean difference between women and men) among White British and the ethnic gap (mean difference between ethnic minorities and White British) among men increased between 2017-2019 and April 2020 (**Figure 2, panel B**)^4^. The increase in the mental health gender gap documented one year ago is not temporary, but persists in March 2021 (**Figure 2, panels A and B**). The point estimates of the ethnic gap among men are similar over time, suggesting a persistent deterioration, albeit the ethnic gap between 2017-2019 and March 2021 is not statistically significant (**Figure 2, panels A and B**).

**Figure 1.**
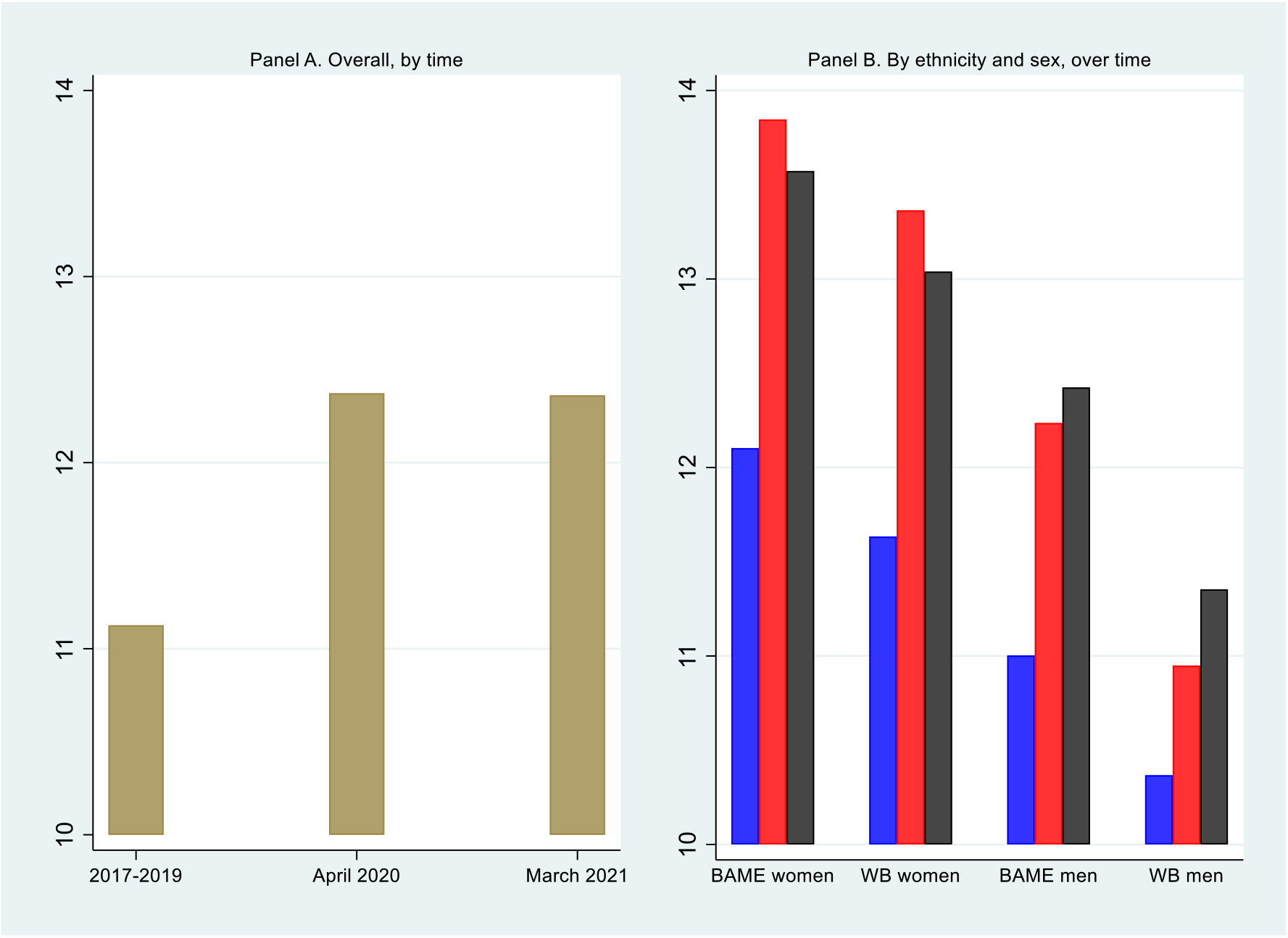
Mental health in the UK in 2017-2019, April 2020 and March 2021: overall and by ethnicity and sex. Note: Secondary data come from UKHLS. Same respondents interviewed in 2017-2019, April 2020 and March 2021. The panels display the mean GHQ-12 score (0-36), overall (panel A) and by ethnicity and sex (panel B), over time (blue: 2017-2019, red: April 2020, dark: March 2021). BAME (Black, Asian and other ethnic minorities), WB (White British).

**Figure 2.**
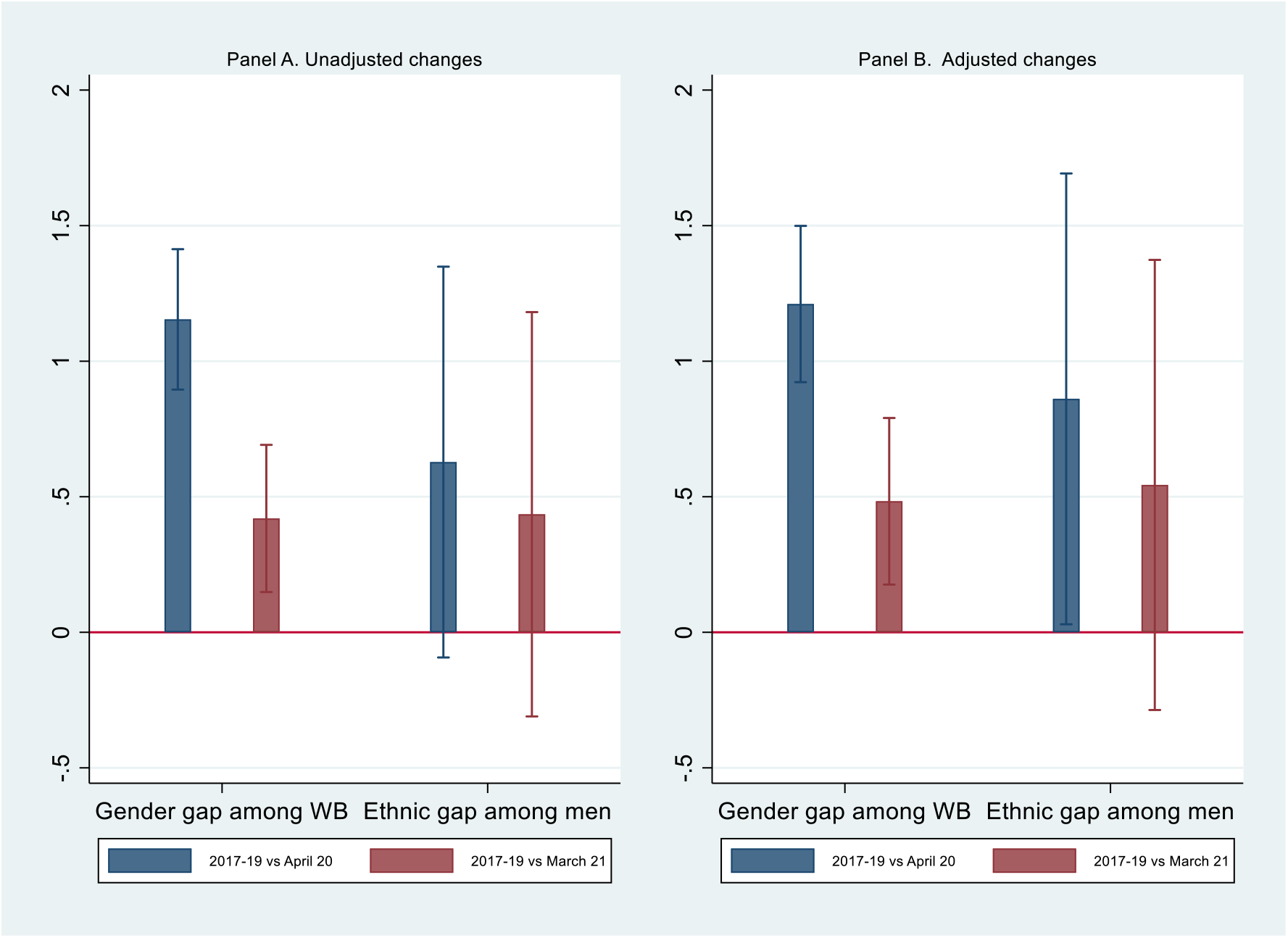
Changes in mental health gaps by ethnicity and sex. Note: Secondary data come from UKHLS. Same respondents interviewed in 2017-2019, April 2020 and March 2021. The panels display regression coefficients (Supplement) capturing the mean change in the GHQ-12 score (−36, 36) between 2017-2019 and April 2020, and between 2017-2019 and March 2021, without (panel A) and with controls (panel B). The controls are described in the Supplement. Each line denotes the corresponding 95% confidence interval.

## Discussion

In contrast to a recent study using Understanding Society, which combines data from the pre-pandemic and an earlier pandemic period (April to October 2020)^6^, we do not find evidence that the level of mental health goes back to pre-pandemic levels. Two reasons might explain this discrepancy of the findings between the two studies. First, we compare the deterioration in mental health between 2017-2019 and late April 2020 with that occurring between 2017-2019 and late March 2021, investigating a longer period of time than previous research^6^. Second, we compare similar seasons (both before summer), while previous research compared different seasons (before vs. after summer 2020).

In terms of inequalities, while the gender gap in mental health deterioration among White British is closing, there is no clear evidence that the ethnic gap among men is changing. Study limitations include: relying on self-reported mental health measures, attrition, and small sample size of individuals from ethnic minorities.

## Data Availability

This short communication uses data from Understanding Society (Wave 9, April 2020 COVID-19 Study, and March 2021 COVID-19 Study). Understanding Society is an initiative funded by the Economic and Social Research Council and various Government Departments, with scientific leadership by the Institute for Social and Economic Research, University of Essex, and survey delivery by NatCen Social Research and Kantar Public. The research data are distributed by the UK Data Service.7,8 Researchers who would like to use Understanding Society need to register with the UK Data Service before being allowed to apply for or download datasets. More information: https://www.understandingsociety.ac.uk/documentation/access-data. The code to replicate the analysis in this short communication will be publicly available upon publication.

## Acknowledgements

This short communication uses data from Understanding Society (Wave 9, April 2020 COVID-19 Study, and March 2021 COVID-19 Study). Understanding Society is an initiative funded by the Economic and Social Research Council and various Government Departments, with scientific leadership by the Institute for Social and Economic Research, University of Essex, and survey delivery by NatCen Social Research and Kantar Public. The research data are distributed by the UK Data Service.^7,8^ Researchers who would like to use Understanding Society need to register with the UK Data Service before being allowed to apply for or download datasets. More information: https://www.understandingsociety.ac.uk/documentation/access-data. The code to replicate the analysis in this short communication will be publicly available upon publication.

## References

1. Banks, J., & Xu, X. (2020). The Mental Health Effects of the First Two Months of Lockdown during the COVID-19 Pandemic in the UK. Volume 41, Issue 3, Fiscal Studies, Special Issue: The COVID-19 Economic Crisis, September, 685–708. https://doi.org/10.1111/1475-5890.12239

2. Daly, M., Sutin, A., & Robinson, E. (2020). Longitudinal changes in mental health and the COVID-19 pandemic: Evidence from the UK Household Longitudinal Study. Psychological Medicine, 1–10. https://doi.org/10.1017/S0033291720004432

3. Pierce, M., Hope, H., Ford, T., Hatch, S., Hotopf, M., John, A., Kontopantelis, E., Webb, R., Wessely, S., McManus, S., & Abel, K. (2020). Mental health before and during the COVID-19 pandemic: a longitudinal probability sample survey of the UK population. The Lancet Psychiatry, Volume 7, Issue 10, Pages 883–892. https://doi.org/10.1016/S2215-0366(20)30308-4

4. Proto, E., & Quintana-Domeque, C. (2021). COVID-19 and mental health deterioration by ethnicity and gender in the UK. PLOS ONE, 16(1): e0244419. https://doi.org/10.1371/journal.pone.0244419

5. Goldberg, D., & Williams, P. (1988) A User’s Guide to the General Health Questionnaire. Windsor: NFER-Nelson.

6. Pierce, M., McManus, S., Hope, H., Hotopf, M., Ford, T., Hatch, S., Hotopf, M., John, A., Kontopantelis, E., Webb, R., Wessely, S., & Abel, K. (2021). Mental health responses to the COVID-19 pandemic: a latent class trajectory analysis using longitudinal UK data. The Lancet Psychiatry, Volume 8, Issue 7, Pages 610–619, https://doi.org/10.1016/S2215-0366(21)00151-6

7. University of Essex, Institute for Social and Economic Research, NatCen Social Research, Kantar Public. (2020). Understanding Society: Waves 1-10, 2009-2019 and Harmonised BHPS: Waves 1-18, 1991-2009. [data collection]. 13th Edition. UK Data Service. SN: 6614, http://doi.org/10.5255/UKDA-SN-6614-14.

8. University of Essex, Institute for Social and Economic Research. (2020). Understanding Society: COVID-19 Study, 2020. [data collection]. 4th Edition. UK Data Service. SN: 8644, http://doi.org/10.5255/UKDA-SN-8644-4.

