## Supplementary material for "On the persistence of mental health deterioration during the COVID-19 pandemic by sex and ethnicity in the UK: evidence from Understanding Society": Online Supplement

This document contains

- **S1 Ethics.** Additional details on Ethical approval statement.
- **S2 Methods.** Additional details on methodology.
- **S3 Table S1.** OLS (1) and Logistic (2) regressions of attrition.
- **S4 Table S2.** OLS regressions of the difference in GHQ-12 between 2017-2019 and April 2020 / March 2021.

**S1 Ethics.** Additional details on Ethical approval statement.

The University of Essex Ethics Committee has approved all data collection on Understanding Society main study and innovation panel waves, including asking consent for all data linkages except to health records. Requesting consent for health record linkage was approved at Wave 1 by the National Research Ethics Service (NRES) Oxfordshire REC A (08/H0604/124), at BHPS Wave 18 by the NRES Royal Free Hospital & Medical School (08/H0720/60) and at Wave 4 by NRES Southampton REC A (11/SC/0274). Approval for the collection of biosocial data by trained nurses in Waves 2 and 3 of the main survey was obtained from the National Research Ethics Service (Understanding Society - UK Household Longitudinal Study: A Biosocial Component, Oxfordshire A REC, Reference: 10/H0604/2).

Ethics approval was granted by the University of Essex Ethics Committee for the COVID-19 web and telephone surveys (ETH1920-1271), for the youth self-completion survey carried out in July (ETH1920-1751), the November youth self-completion survey (ETH2021-0279), and the March 2021 youth self-completion survey (ETH2021-0969). The March 2021 web survey was reviewed, and ethics approval granted by the NHS Health Research Authority, London – City & East Research Ethics Committee (reference 21/HRA/0644).

**S2 Methods.** Additional Information on Methodology

Sampling design features (i.e., weighting and adjusting standard errors for primary sampling units and strata) are accounted for by using the *svyset* command in Stata:[1]

```
svyset psu [pweight=indinui_xw_all], strata(strata) singleunit(centered)
```

As reported in the short communication the attrition rate was 25.7%. Men, ethnic minorities, individuals with a higher GHQ-12 score and poorer individuals are more likely to be attritors (**Table S1**).

To estimate the changes in mental health by sex and ethnicity, we perform two types of regressions, without controls and with controls, following a previous published study.[2] The list of control variables is defined in that previous study. **Figure 2** in the short communication plots the estimated coefficients on the sex (women) and ethnicity (BAME) variables from **Table S2**.

**S3 Table S1.** OLS (1) and Logistic (2) regressions of attrition from April 2020 to March 2021.

The table reports the coefficients (and the 95% confidence interval) for the OLS regression and Odds Ratios (OR) for the Logistic regression.

| | Dependent variable | |
|---|---|---|
| | (1) Attrition | (2) Attrition |
| Female (1 if woman, 0 if man) | -0.023*** | 0.883*** |
| | (-0.041, -0.006) | (0.804, 0.969) |
| BAME (1 if ethnic minority, 0 if White British) | 0.131*** | 1.861*** |
| | (0.097, 0.166) | (1.600, 2.163) |
| GHQ-12 (0-36) | 0.004*** | 1.020*** |
| | (0.002, 0.006) | (1.011, 1.029) |
| Net personal income (£1K) | -0.004 | 0.976 |
| | (-0.009, 0.001) | (0.944, 1.009) |
| Observations | 14,382 | 14,382 |
| R-squared | 0.010 | -- |

*Note: Attrition = 1 if individual is observed in 2017-2019, April 2020 and March 2021; = 0 if individual observed in 2017-2019 and April 2020 but not in March 2021. Each regression includes the variables Female, BAME, GHQ-12, Net personal income (£1K) and a constant term. 95% confidence intervals in parentheses.*
**** p<0.01*

**S4 Table S2. OLS regressions of the difference in GHQ-12 between 2017-2019 and April 2020 / March 2021.**

The table reports the coefficients (and the 95% confidence interval) for the OLS regressions.

| | Dependent variable | | | |
|---|---|---|---|---|
| | (1) Difference in GHQ-12 between 2017-2019 and April 2020 | (2) Difference in GHQ-12 between 2017-2019 and March 2021 | (3) Difference in GHQ-12 between 2017-2019 and April 2020 | (4) Difference in GHQ-12 between 2017-2019 and March 2021 |
| Female | 1.154*** | 0.420*** | 1.211*** | 0.483*** |
| | (0.895, 1.413) | (0.148, 0.691) | (0.923, 1.499) | (0.176, 0.791) |
| BAME | 0.628 | 0.435 | 0.861** | 0.543 |
| | (-0.093, 1.349) | (-0.310, 1.181) | (0.030, 1.692) | (-0.287, 1.374) |
| Female × BAME | -0.694 | -0.371 | -0.937 | -0.676 |
| | (-1.811, 0.424) | (-1.364, 0.621) | (-2.128, 0.254) | (-1.726, 0.375) |
| Age $\leq$ 24 | | | 2.355*** | 1.594** |
| | | | (0.930, 3.780) | (0.370, 2.819) |
| Age 25-34 | | | 1.804*** | 1.350*** |
| | | | (1.025, 2.583) | (0.574, 2.127) |
| Age 35-44 | | | 0.774** | 0.964*** |
| | | | (0.024, 1.524) | (0.294, 1.633) |
| Age 45-54 | | | 0.145 | 0.541 |
| | | | (-0.419, 0.710) | (-0.015, 1.097) |
| Age 55-64 | | | 0.289 | 0.235 |
| | | | (-0.182, 0.759) | (-0.220, 0.691) |
| Living with a partner | | | -0.176 | -0.244 |
| | | | (-0.603, 0.250) | (-0.649, 0.161) |
| January | | | 0.085 | 0.113 |
| | | | (-0.651, 0.821) | (-0.641, 0.867) |
| February | | | 0.057 | -0.081 |
| | | | (-0.768, 0.882) | (-0.893, 0.730) |
| March | | | -0.075 | -0.121 |
| | | | (-0.849, 0.698) | (-0.888, 0.647) |
| May | | | 0.239 | -0.068 |
| | | | (-0.535, 1.014) | (-0.842, 0.706) |
| June | | | 0.149 | 0.425 |
| | | | (-0.626, 0.923) | (-0.353, 1.203) |
| July | | | 0.222 | 0.104 |
| | | | (-0.540, 0.984) | (-0.663, 0.872) |

| | | |
|---|---|---|
| August | 0.235 | 0.434 |
| | (-0.471, 0.940) | (-0.334, 1.202) |
| September | 0.762 | 0.625 |
| | (-0.014, 1.538) | (-0.182, 1.432) |
| October | -0.100 | 0.004 |
| | (-0.890, 0.690) | (-0.793, 0.802) |
| November | -0.158 | 0.029 |
| | (-0.930, 0.615) | (-0.725, 0.782) |
| December | -0.281 | -0.506 |
| | (-1.050, 0.488) | (-1.283, 0.271) |
| Face-to-face | 0.292 | 0.439** |
| | (-0.052, 0.636) | (0.090, 0.789) |
| Household size | -0.003 | 0.025 |
| | (-0.166, 0.159) | (-0.132, 0.182) |
| London | 0.447 | 0.479 |
| | (-0.255, 1.150) | (-0.123, 1.080) |
| Wales | 0.300 | 0.147 |
| | (-0.444, 1.044) | (-0.656, 0.951) |
| Scotland | -0.071 | -0.174 |
| | (-0.645, 0.504) | (-0.835, 0.487) |
| Northern Ireland | -0.013 | -0.262 |
| | (-1.328, 1.302) | (-1.627, 1.103) |
| BA or higher | 0.005 | -0.136 |
| | (-0.441, 0.450) | (-0.551, 0.280) |
| Diploma or equivalent | -0.076 | -0.413 |
| | (-0.643, 0.490) | (-0.966, 0.140) |
| A Level or equivalent | -0.067 | -0.272 |
| | (-0.641, 0.506) | (-0.866, 0.323) |
| GCSE or equivalent | -0.170 | -0.243 |
| | (-0.620, 0.280) | (-0.686, 0.200) |
| Self-employed | 0.513 | 0.427 |
| | (-0.120, 1.146) | (-0.101, 0.956) |
| Unemployed | -2.085*** | -1.923*** |
| | (-3.240, -0.931) | (-3.185, -0.660) |
| Retired | 0.923*** | 1.185*** |
| | (0.458, 1.388) | (0.742, 1.628) |
| Family care or home | -0.650 | -1.258** |
| | (-1.833, 0.533) | (-2.354, -0.163) |
| Student | -0.982 | -0.860 |
| | (-2.482, 0.517) | (-2.215, - 0.495) |
| Disabled | -2.756*** | -1.970*** |
| | (-4.065, -1.447) | (-3.346, -0.595) |
| Other | 1.045 | 0.688 |
| | (-0.756, 2.847) | (-1.080, 2.456) |
| Net personal income (£1K) | 0.026 | 0.015 |
| | (-0.076, 0.129) | (-0.075, 0.104) |
| Health conditions | -0.128 | 0.101 |

| | | | | |
|---|---|---|---|---|
| | | | (-0.467, 0.212) | (-0.214, 0.416) |
| Observations | 10,445 | 10,445 | 9,045 | 9,045 |
| R-squared | 0.009 | 0.001 | 0.033 | 0.019 |

*Note: 95% confidence intervals in parentheses.*
**** p<0.01, ** p<0.05*

**References**


1. Understanding Society COVID-19, USER GUIDE, Version 8.0, May 2021, https://www.understandingsociety.ac.uk/sites/default/files/downloads/documentation/covid-19/user-guides/covid-19-user-guide.pdf

2. Proto, E., & Quintana-Domeque, C. (2021). COVID-19 and mental health deterioration by ethnicity and gender in the UK. PLOS ONE, 16(1): e0244419. https://doi.org/10.1371/journal.pone.0244419
